# Cause-specific mortality among banana plantation workers in the French West Indies

**DOI:** 10.1101/2025.04.30.25326769

**Authors:** Danièle Luce, Juliette Gambaretti, Léah Michineau, Luc Multigner, Christine Barul

## Abstract

**Objective:** To describe cause-specific mortality patterns of banana plantation workers in the French West Indies.

**Methods:** The study included 11221 farmers and farm workers who had work in banana cultivation in the French West indies (Guadeloupe or Martinique) between 1973 and 1993, followed up from January 1981 to December 2017. We calculated standardized mortality ratios (SMRs), causal mortality ratios (CMRs) and relative standardized mortality ratios (rSMRs) using regional reference rates.

**Results:** SMR analyses showed mortality deficits in overall mortality and for almost all causes of deaths. In contrast, analyses using CMRs revealed a significant excess in overall mortality. The CMRs were significantly elevated for all cancers combined and for stomach cancer, colorectal cancer, prostate cancer and hematopoietic malignancies, as well as for several non-cancer causes of death, including diabetes mellitus, Parkinson’s disease, Alzheimer’s disease, non-ischemic heart diseases, pneumonia and diseases of the skin and subcutaneous tissue. rSMRs were in general consistent with CMRs.

**Conclusion:** The CMR approach showed an elevated mortality for several causes of death, for which work in banana farming and/or exposure to pesticides are plausible explanations.

## Introduction

In the French West Indies, banana cultivation has long been an important part of the regional agriculture. However, banana plantation workers face numerous occupational hazards. As in any tropical environment, heavy parasitic pressure leads to intensive use of pesticides. In particular, the use of chlordecone, a persistent organochlorine insecticide widely used in the French West Indies from the 1970s to the early 1990s to control banana weevil, has raised major public health concerns due to its persistence in the environment and its health impacts [1]. Alongside exposure to chlordecone, banana workers are exposed to many other pesticides. They are also subjected to physical, biological and biomechanical hazards.

We set up a historical cohort of banana workers potentially exposed to chlordecone in the French West Indies. Although the focus was initially on the effects of chlordecone exposure, this cohort also provides an opportunity to study long-term health effects in this population facing unique occupational and environmental risks. We previously reported a first mortality analysis of this cohort for the period 2000-2015 [2]. Other epidemiological data on mortality patterns specific to banana workers remain limited to a single cohort in Costa-Rica [3]. Here we aim to describe the mortality of banana workers in our cohort with a follow-up extended to 1981-2017, by examining cause-specific mortality for various diseases, using three complementary analytical approaches.

## Methods

### Population study and follow-up

Cohort construction has been described in details previously [2]. Briefly, the cohort includes farm owners and farm workers, who had worked on a banana plantation in the French West Indies (Guadeloupe or Martinique) between 1973 and 1993, the period of authorized chlordecone use. The cohort was assembled retrospectively from social security fund records and general agricultural censuses. Vital status was obtained for the period 1973-2017 by linkage with the French national vital status registry. Causes of death were obtained from the French national mortality registry (Centre d’épidémiologie sur les causes médicales de décès, CepiDC). For French overseas regions, deaths occurring before 2000 were collected but not included in this registry, because they had not been subject to standardized validation procedures. To fill this gap, the CepiDC carried out a comprehensive validation process to provide us with cause-specific mortality data for the period 1981-1999 for the cohort and for the general population of Guadeloupe and Martinique.

The initial cohort comprised 13417 workers, of whom 13213 were still alive on January 1, 1981. We excluded 66 workers for whom the cause of death could not be determined, as well as 357 workers born before 1910 for data quality issues. Workers with a missing date of start of employment in banana farming were also excluded from all analyses, which finally included 11221 workers (8825 men, 2396 women; 6138 farm owners, 5083 farm workers; 3591 in Guadeloupe, 7630 in Martinique).

For each worker, the follow-up began on January 1, 1981, or the date on which employment in banana farming started, whichever occurred later, and ended on December 31, 2017, or the date of death, whichever occurred first. The 11221 workers included in mortality analyses contributed 321,573 person-years of observation, with an average follow-up of 28,7 years.

### Statistical analysis

Cohort mortality was compared with that of the general population of Guadeloupe and Martinique by calculating standardized mortality ratios (SMRs), controlling for region, gender, age (15-24, 25-34, … 85 and over) and period (1981-1983, 1984-1987, … 2016-2017). We also computed causal mortality ratios (CMRs), a measure considered as more reliable than the SMR [4–6]. Indeed, the classical SMR calculation uses the observed person-times accrued in the cohort to calculate the expected number of deaths, assuming that exposure (here, work in banana cultivation) does not affect the person-time distribution in the cohort. The CMR does not require this assumption, as it uses the person-time that would be expected if the cohort had actually experienced the mortality rates of the reference population, by taking into account the survival probability during the follow-up. For SMRs and CMRs, we computed exact 95% confidence intervals (CI) based on the Poisson distribution.

To account for the healthy worker effect typically observed in occupational cohorts, we also calculated a relative SMR (rSMR) for each cause of death of interest, defined as the ratio of the SMR for a given cause to the SMR for all other causes, excluding the cause of interest [5, 6]. The rSMR may reduce the healthy worker effect, assuming comparable bias for all causes.

SMRs, CMRs and rSMRs were computed for the entire cohort and were also calculated by gender, by employment status (farm owners and farm workers), and by region (Guadeloupe and Martinique).

The analyses only considered the underlying cause of death, as the contributing causes were not available for most deaths that occurred between 1981 and 1999. Causes of death were grouped according to the categories of the European short list for causes of death, which considers the change in classification between 1981-1999 (International Classification of Diseases, 9th Revision, ICD-9) and 2000-2017 (10th Revision, ICD-10). The causes of death studied and their ICD codes are listed in Supplementary Table 1.

## Results

Table 1 shows the SMRs, CMRs and rSMRs for the various causes of death in the entire cohort.

**Table 1.** Standardized mortality ratios (SMRs), causal mortality ratios (CMRs) and relative SMRs (rSMRs) among the French West Indies cohort of banana workers for the period 1981-2017.

|  | Observed<br>deaths | Expected<br>deaths<br>(SMR) | SMR (95% CI) | Expected<br>deaths<br>(CMR) | CMR (95% CI) | rSMR (95% CI) |
| --- | --- | --- | --- | --- | --- | --- |
| All causes of death | 5165 | 6196 | 0.83 (0.81-0.86) | 3950 | 1.31 (1.27-1.34) |  |
| Infectious and parasitic diseases | 127 | 194 | 0.66 (0.55-0.78) | 129 | 0.99 (0.82-1.17) | 0.78 (0.65-0.93) |
| All cancers | 1392 | 1594 | 0.87 (0.83-0.92) | 1064 | 1.31 (1.24-1.38) | 1.06 (1.00-1.13) |
| Lip, buccal cavity, pharynx | 70 | 75 | 0.94 (0.73-1.18) | 60 | 1.17 (0.91-1.48) | 1.12 (0.87-1.42) |
| Esophagus | 51 | 63 | 0.81 (0.60-1.06) | 49 | 1.03 (0.77-1.36) | 0.97 (0.72-1.27) |
| Stomach | 174 | 159 | 1.09 (0.94-1.27) | 110 | 1.59 (1.36-1.84) | 1.32 (1.13-1.54) |
| Colon, rectum and anus | 94 | 110 | 0.86 (0.69-1.05) | 72 | 1.31 (1.06-1.60) | 1.03 (0.83-1.26) |
| Liver and intrahepatic bile duct | 33 | 63 | 0.52 (0.36-0.74) | 46 | 0.71 (0.49-1.00) | 0.63 (0.43-0.88) |
| Pancreas | 61 | 80 | 0.77 (0.59-0.98) | 54 | 1.12 (0.86-1.44) | 0.92 (0.70-1.18) |
| Larynx | 19 | 27 | 0.72 (0.43-1.12) | 20 | 0.95 (0.57-1.48) | 0.86 (0.52-1.35) |
| Lung, trachea and bronchus | 106 | 137 | 0.78 (0.64-0.94) | 99 | 1.07 (0.88-1.30) | 0.93 (0.76-1.13) |
| Skin | 4 | 7 | 0.57 (0.16-1.47) | 5 | 0.78 (0.21-2.01) | 0.69 (0.19-1.76) |
| Breast | 27 | 33 | 0.82 (0.54-1.20) | 27 | 1.02 (0.67-1.48) | 0.99 (0.65-1.44) |
| Uterus | 27 | 26 | 1.02 (0.67-1.49) | 21 | 1.26 (0.83-1.84) | 1.23 (0.81-1.79) |
| Ovary | 5 | 8 | 0.61 (0.20-1.42) | 7 | 0.75 (0.24-1.74) | 0.73 (0.24-1.70) |
| Prostate | 372 | 397 | 0.94 (0.84-1.04) | 216 | 1.72 (1.55-1.90) | 1.13 (1.02-1.26) |
| Kidney | 4 | 12 | 0.34 (0.09-0.88) | 9 | 0.45 (0.12-1.16) | 0.41 (0.11-1.05) |
| Bladder | 26 | 31 | 0.84 (0.55-1.23) | 20 | 1.32 (0.86-1.93) | 1.01 (0.66-1.48) |
| Brain and central nervous system | 12 | 17 | 0.70 (0.36-1.22) | 14 | 0.87 (0.45-1.52) | 0.84 (0.43-1.46) |
| Lymphatic and hematopoietic cancer | 116 | 133 | 0.87 (0.72-1.05) | 90 | 1.29 (1.07-1.55) | 1.05 (0.86-1.26) |
| Diseases of the blood and of blood-forming organs | 17 | 26 | 0.66 (0.38-1.05) | 17 | 0.98 (0.57-1.57) | 0.79 (0.46-1.26) |
| Endocrine, nutritional and metabolic disease | 287 | 343 | 0.84 (0.74-0.94) | 206 | 1.39 (1.24-1.56) | 1.01 (0.89-1.13) |
| Diabetes mellitus | 216 | 240 | 0.90 (0.78-1.03) | 150 | 1.44 (1.26-1.65) | 1.08 (0.94-1.24) |
| Mental and behavioral disorders | 148 | 196 | 0.75 (0.64-0.89) | 139 | 1.07 (0.90-1.25) | 0.90 (0.76-1.06) |
| Alcohol abuse | 95 | 113 | 0.84 (0.68-1.03) | 96 | 0.99 (0.80-1.22) | 1.01 (0.81-1.23) |
| Diseases of the nervous system | 280 | 295 | 0.95 (0.84-1.07) | 172 | 1.63 (1.44-1.83) | 1.15 (1.01-1.29) |
| Parkinson's disease | 47 | 58 | 0.81 (0.60-1.08) | 32 | 1.45 (1.07-1.93) | 0.97 (0.71-1.30) |
| Alzheimer's disease | 114 | 115 | 0.99 (0.82-1.19) | 60 | 1.92 (1.58-2.30) | 1.19 (0.98-1.44) |
| Diseases of the circulatory system | 1515 | 1792 | 0.85 (0.80-0.89) | 1122 | 1.35 (1.28-1.42) | 1.02 (0.96-1.08) |
| Ischemic heart diseases | 184 | 239 | 0.77 (0.66-0.89) | 160 | 1.15 (0.99-1.33) | 0.92 (0.79-1.07) |
| Other heart diseases | 381 | 459 | 0.83 (0.75-0.92) | 278 | 1.37 (1.24-1.52) | 1.00 (0.90-1.11) |
| Cerebrovascular diseases | 408 | 687 | 0.59 (0.54-0.65) | 442 | 0.92 (0.84-1.02) | 0.69 (0.62-0.76) |
| Diseases of the respiratory system | 243 | 333 | 0.73 (0.64-0.83) | 187 | 1.30 (1.14-1.47) | 0.87 (0.76-0.99) |
| Pneumonia | 101 | 137 | 0.74 (0.60-0.90) | 73 | 1.39 (1.13-1.69) | 0.88 (0.72-1.08) |
| Chronic lower respiratory diseases | 67 | 88 | 0.76 (0.59-0.97) | 57 | 1.17 (0.90-1.48) | 0.91 (0.71-1.16) |
| Diseases of the digestive system | 252 | 304 | 0.83 (0.73-0.94) | 213 | 1.19 (1.04-1.34) | 0.99 (0.87-1.13) |
| Cirrhosis, fibrosis and chronic hepatitis | 87 | 99 | 0.88 (0.70-1.09) | 82 | 1.07 (0.85-1.32) | 1.06 (0.84-1.31) |
| Diseases of the skin and subcutaneous tissues | 31 | 30 | 1.02 (0.70-1.45) | 18 | 1.74 (1.18-2.47) | 1.23 (0.84-1.75) |
| Diseases of the musculoskeletal system/connective tissue | 18 | 24 | 0.76 (0.45-1.20) | 15 | 1.21 (0.72-1.91) | 0.91 (0.54-1.44) |
| Diseases of the genitourinary system | 103 | 129 | 0.80 (0.65-0.97) | 78 | 1.33 (1.08-1.61) | 0.96 (0.78-1.17) |
| Diseases of the kidney and ureter | 64 | 84 | 0.76 (0.59-0.97) | 53 | 1.20 (0.93-1.54) | 0.91 (0.70-1.17) |
| Symptoms, signs, ill-defined causes | 401 | 503 | 0.80 (0.72-0.88) | 270 | 1.49 (1.35-1.64) | 0.95 (0.86-1.06) |
| External causes | 303 | 379 | 0.80 (0.71-0.89) | 287 | 1.06 (0.94-1.18) | 0.96 (0.85-1.07) |

SMRs analyses indicated that all-cause mortality was significantly lower in the cohort than in the general population. The same was true for cancer mortality, and for almost all other major causes of death. For most cancer sites, SMRs were below 1, except for stomach cancer for which a slight non-significant excess of deaths was observed. No SMR was significantly greater than 1, whatever the cause of death.

In contrast, the CMRs results revealed a significant excess in overall mortality. The CMRs were significantly elevated for all cancers combined and for stomach cancer, colorectal cancer, prostate cancer and hematopoietic malignancies. CMRs significantly greater than 1 were also observed for a number of non-cancer causes of death, including diabetes mellitus, Parkinson’s disease, Alzheimer’s disease, non-ischemic heart diseases, pneumonia and diseases of the skin and subcutaneous tissue. The results of rSMRs were in general consistent with those of CMRs with regard to the direction of the association, although rSMRs were generally lower than CMRs.

Separate analyses by gender, employment status and region are reported in Supplementary tables 2, 3 and 4 respectively. No substantial differences were found.

## Discussion

SMR analyses showed mortality deficits, as in the previous analysis covering the period 2000- 2015 [2]. The inclusion of deaths from the 1981-1999 period led to lower SMRs for most causes of death, due to a more pronounced healthy worker effect for this period.

Conversely, CMRs findings and, to a lesser extent, rSMRs findings demonstrated an increase in mortality from several causes. These estimates are likely to be less biased than the SMR. These results are overall consistent with previous studies and agricultural exposures are plausible explanations for most of the elevated CMRs. Elevated risks of prostate cancer, stomach cancer and hematopoietic malignancies have been consistently observed in cohorts of agricultural workers [7]. There is evidence that the use of pesticides increases the risk of Parkinson’s disease and Alzheimer’s disease [8]. Diabetes mellitus has been found to be associated with exposure to pesticides, particularly insecticides [9]. Epidemiological data on the role of pesticides in the occurrence of colorectal cancer are somewhat conflicting, but positive associations with certain pesticides or tasks have been reported in several studies [10, 11]. We observed a decrease in mortality from cerebrovascular diseases, but a significantly increased CMR for non-ischemic heart diseases. Cardiovascular mortality is generally decreased in agricultural workers, although elevated CMRs for several cardiovascular causes were reported in the Agricultural health study [5]. Increased mortality from pneumonia and from skin diseases were rarely reported and may be related to biological hazards, limited access to healthcare, or may be chance findings.

This study confirms the value, previously highlighted [5], of using other approaches than the classical SMR to analyze mortality in agricultural populations. The long follow-up is another strength of our study. The limitations are those inherent to mortality studies, notably the lack of accuracy of death certificates in identifying some diseases, compounded here by the lower reliability of mortality data prior to the year 2000. Other limitations include the inability to attribute the observed excess deaths to specific occupational exposures, the lack of data on lifestyle confounders and the healthy worker effect, though rSMR mitigates this latter bias.

We have provided a general description of mortality patterns in this cohort of banana workers. Subsequent analyses will examine the associations between occupational exposure to pesticides, assessed by a specific a crop-exposure matrix, and cause-specific mortality.

## Acknowledgments

We are grateful to the General Social Security Funds (Caisses Générales de Sécurité Sociale) of Guadeloupe and Martinique for their collaboration. We warmly thank Amandine Vaidie and Julien Dugas for their contribution to the cohort set-up and data management; we also thank Marie Barrau, Alexandra Doens, Sylvia Janky, Vanessa Jupiter, Jessica Obertan, Stéphane Renaud, Nathalie Surville-Barland for their help in data collection. We also would like to thank the Centre d’épidémiologie sur les causes médicales de décès, and in particular Walid Ghosn for their work on the causes of death in French overseas territories for the period before 2000.

## Compliance with ethical standards

The study was approved by the Institutional Review Board of the French National Institute of Health and Medical Research (IRB-INSERM, n° 15-193), and by the French Data Protection Authority (CNIL n° DR-215-200). Access to the nominative questionnaires of the agricultural censuses was authorized by the Committee on Statistical Confidentiality of the National Institute of Statistics and Economic Studies (session 14 January 2015, n°E270).

## Data availability statement

Data are not publicly available due to legal restrictions. Deidentified data are available upon reasonable request to the corresponding author.

## References

1. Multigner L, Kadhel P, Rouget F, Blanchet P, Cordier S. Chlordecone exposure and adverse effects in French West Indies populations. Environ Sci Pollut Res Int. 2016;23:3–8.

2. Luce D, Dugas J, Vaidie A, Michineau L, El-Yamani M, Multigner L. A cohort study of banana plantation workers in the French West Indies: first mortality analysis (2000-2015). Environ Sci Pollut Res Int. 2020;27:41014–22.

3. Hofmann J, Guardado J, Keifer M, Wesseling C. Mortality among a cohort of banana plantation workers in Costa Rica. Int J Occup Environ Health. 2006;12:321–8.

4. Richardson DB, Keil AP, Cole SR, MacLehose RF. Observed and Expected Mortality in Cohort Studies. Am J Epidemiol. 2017;185:479–86.

5. Shrestha S, Parks CG, Keil AP, Umbach DM, Lerro CC, Lynch CF, et al. Overall and cause-specific mortality in a cohort of farmers and their spouses. Occup Environ Med. 2019;76:632–43.

6. Bovio N, Richardson DB, Guseva Canu I. Sex-specific risks and trends in lung cancer mortality across occupations and economic activities in Switzerland (1990-2014). Occup Environ Med. 2020;77:540–8.

7. Blair A, Freeman LB. Epidemiologic Studies of Cancer in Agricultural Populations: Observations and Future Directions. J Agromedicine. 2009;14:125–31.

8. Gunnarsson L-G, Bodin L. Occupational Exposures and Neurodegenerative Diseases—A Systematic Literature Review and Meta-Analyses. Int J Environ Res Public Health. 2019;16:337.

9. Wei Y, Wang L, Liu J. The diabetogenic effects of pesticides: Evidence based on epidemiological and toxicological studies. Environmental Pollution. 2023;331:121927.

10. Matich EK, Laryea JA, Seely KA, Stahr S, Su LJ, Hsu P-C. Association between pesticide exposure and colorectal cancer risk and incidence: A systematic review. Ecotoxicol Environ Saf. 2021;219:112327.

11. Xie P-P, Zong Z-Q, Qiao J-C, Li Z-Y, Hu C-Y. Exposure to pesticides and risk of colorectal cancer: A systematic review and meta-analysis. Environ Pollut. 2024;345:123530.

