## Supplementary Tables 1-4 for "Cause-specific mortality among banana plantation workers in the French West Indies"

**Supplementary table 1.** Causes of death and their codes in the International Classification of diseases 9th revision (ICD 9, 1981-2000) and 10th revision (ICD 10, 2000-2017)

| | ICD 10 | ICD 9 |
|---|---|---|
| All causes of death | A00-Y89 | 001-E999 |
| Infectious and parasitic diseases | A00-B99 | 001-139 |
| All cancers | C00-C97 | 140-208 |
| Lip, buccal cavity, pharynx | C00-C14 | 140-149 |
| Esophagus | C15 | 150 |
| Stomach | C16 | 151 |
| Colon, rectum and anus | C18-C21 | 153-154 |
| Liver and intrahepatic bile duct | C22 | 155 |
| Pancreas | C25 | 157 |
| Larynx | C32 | 161 |
| Lung, trachea and bronchus | C33-C34 | 162 |
| Skin | C43 | 172 |
| Breast | C50 | 174-175 |
| Uterus | C53-C55 | 179,180,18 |
| Ovary | C56 | 183.0 |
| Prostate | C61 | 185 |
| Kidney | C64 | 189.0 |
| Bladder | C67 | 188 |
| Brain and central nervous system | C70-C72 | 191-192 |
| Lymphatic and hematopoietic tissues | C81-C96 | 200-208 |
| Diseases of the blood and of blood-forming organs | D50-D89 | 279-289 |
| Endocrine, nutritional and metabolic disease | E00-E90 | 240-279 |
| Diabetes mellitus | E10-E14 | 250 |
| Mental and behavorial disorders | F01-F99 | 290-319 |
| Alcohol abuse | F10 | 291, 303 |
| Diseases of the nervous system | G00-H95 | 320-389 |
| Parkinson's disease | G20 | 332.0 |
| Alzheimer's disease | G30 | 331.0 |
| Diseases of the circulatory system | I00-I99 | 390-459 |
| Ischemic heart diseases | I20-I25 | 410-414 |
| Other heart diseases | I30-I51 | 420-429 |
| Cerebrovascular diseases | I60-I69 | 430-438 |
| Diseases of the respiratory system | J00-J99 | 460-519 |
| Pneumonia | J12-J18 | 480-486 |
| Chronic lower respiratory diseases | J40-J47 | 490-494, |
| Diseases of the digestive system | K00-K93 | 520-579 |
| Cirrhosis, fibrosis and chronic hepatitis | K70, K73-K74 | 571 |
| Diseases of the skin and subcutaneous tissues | L00-L99 | 680-709 |
| Diseases of the musculoskeletal system/connective tissue | M00-M99 | 710-739 |
| Diseases of the genitourinary system | N00-N99 | 580-629 |
| Diseases of the kidney and ureter | N00-N29 | 580-594 |
| Symptoms, signs, ill-defined causes | R00-R99 | 780-799 |
| External causes | V01-Y89 | E800-E999 |

**Supplementary table 2**. Standardized mortality ratios (SMRs), causal mortality ratios (CMRs) and relative SMRs (rSMRs) by gender

| | Men | | | | Women | | | |
|---|---|---|---|---|---|---|---|---|
| | **Obs** | **SMR (95% CI)** | **CMR (CI 95%)** | **rSMR (95%CI)** | **Obs** | **SMR (95% CI)** | **CMR (CI 95%)** | **rSMR (95%CI)** |
| All causes of death | 4277 | 0.83 (0.80-0.85) | 1.32 (1.28-1.36) | | 888 | 0.85 (0.80-0.91) | 1.24 (1.16-1.33) | |
| Infectious and parasitic diseases | 102 | 0.63 (0.51-0.76) | 0.96 (0.78-1.16) | 0.75 (0.61-0.92) | 25 | 0.80 (0.52-1.18) | 1.13 (0.73-1.66) | 0.93 (0.60-1.39) |
| All cancers | 1187 | 0.87 (0.82-0.92) | 1.34 (1.26-1.41) | 1.06 (1.00-1.14) | 205 | 0.90 (0.78-1.03) | 1.16 (1.01-1.34) | 1.07 (0.91-1.26) |
| Lip, buccal cavity, pharynx | 70 | 0.96 (0.75-1.21) | 1.21 (0.94-1.52) | 1.16 (0.90-1.47) | 0 | - | - | - |
| Esophagus | 49 | 0.81 (0.60-1.08) | 1.04 (0.77-1.38) | 0.98 (0.72-1.30) | 2 | 0.68 (0.08-2.45) | 0.83 (0.10-3.01) | 0.79 (0.10-2.88) |
| Stomach | 149 | 1.07 (0.90-1.25) | 1.56 (1.32-1.84) | 1.30 (1.09-1.53) | 25 | 1.29 (0.84-1.91) | 1.74 (1.12-2.56) | 1.53 (0.98-2.27) |
| Colon, rectum and anus | 70 | 0.82 (0.64-1.03) | 1.29 (1.01-1.63) | 0.99 (0.77-1.25) | 24 | 0.99 (0.63-1.47) | 1.35 (0.86-2.01) | 1.16 (0.74-1.74) |
| Liver | 28 | 0.52 (0.34-0.75) | 0.71 (0.47-1.03) | 0.62 (0.41-0.90) | 5 | 0.56 (0.18-1.31) | 0.71 (0.23-1.67) | 0.65 (0.21-1.53) |
| Pancreas | 47 | 0.74 (0.54-0.98) | 1.11 (0.82-1.48) | 0.88 (0.65-1.18) | 14 | 0.88 (0.48-1.48) | 1.17 (0.64-1.96) | 1.06 (0.56-1.74) |
| Larynx | 19 | 0.73 (0.44-1.14) | 0.96 (0.58-1.51) | 0.88 (0.53-1.38) | 0 | - | - | - |
| Lung | 91 | 0.74 (0.60-0.91) | 1.03 (0.83-1.27) | 0.89 (0.72-1.10) | 15 | 1.07 (0.60-1.77) | 1.39 (0.78-2.29) | 1.29 (0.70-2.09) |
| Skin | 4 | 0.69 (0.19-1.76) | 0.95 (0.26-2.44) | 0.82 (0.23-2.12) | 0 | - | - | 0.00 (-) |
| Breast | 4 | 1.67 (0.45-4.27) | 2.11 (0.57-5.39) | 2.00 (0.55-5.16) | 23 | 0.76 (0.48-1.13) | 0.93 (0.59-1.40) | 0.91 (0.56-1.33) |
| Uterus | | | | | 27 | 1.03 (0.68-1.49) | 1.26 (0.83-1.84) | 1.23 (0.79-1.77) |
| Ovary | | | | | 5 | 0.61 (0.20-1.42) | 0.75 (0.24-1.74) | 0.73 (0.23-1.66) |
| Prostate | 372 | 0.94 (0.84-1.04) | 1.72 (1.55-1.90) | 1.13 (1.02-1.27) | | | | |
| Kidney | 2 | 0.21 (0.02-0.74) | 0.28 (0.03-1.02) | 0.25 (0.03-0.90) | 2 | 1.01 (0.12-3.66) | 1.25 (0.15-4.52) | 1.22 (0.14-4.30) |
| Bladder | 20 | 0.75 (0.46-1.16) | 1.22 (0.74-1.88) | 0.90 (0.55-1.40) | 6 | 1.37 (0.50-2.98) | 1.82 (0.67-3.96) | 1.64 (0.59-3.51) |
| Brain and central nervous system | 9 | 0.65 (0.30-1.24) | 0.83 (0.38-1.57) | 0.78 (0.36-1.49) | 3 | 0.88 (0.18-2.57) | 1.03 (0.21-3.02) | 1.05 (0.21-3.02) |
| Lymphatic and hematopoietic tissues | 96 | 0.89 (0.72-1.09) | 1.37 (1.11-1.67) | 1.07 (0.87-1.32) | 20 | 0.79 (0.48-1.22) | 1.03 (0.63-1.59) | 0.95 (0.56-1.44) |
| Diseases of the blood and of blood-forming organs | 14 | 0.69 (0.38-1.16) | 1.05 (0.58-1.77) | 0.83 (0.45-1.39) | 3 | 0.54 (0.11-1.58) | 0.75 (0.15-2.19) | 0.65 (0.13-1.86) |
| Endocrine, nutritional and metabolic disease | 215 | 0.82 (0.72-0.94) | 1.41 (1.23-1.62) | 0.99 (0.86-1.14) | 72 | 0.89 (0.69-1.12) | 1.33 (1.04-1.68) | 1.07 (0.81-1.33) |
| Diabetes mellitus | 158 | 0.88 (0.75-1.02) | 1.45 (1.24-1.70) | 1.05 (0.90-1.24) | 58 | 0.97 (0.73-1.25) | 1.41 (1.07-1.82) | 1.16 (0.86-1.49) |

| | Men | | | | Women | | | |
|---|---|---|---|---|---|---|---|---|
| | Obs | SMR (95% CI) | CMR (CI 95%) | rSMR (95%CI) | Obs | SMR (95% CI) | CMR (CI 95%) | rSMR (95%CI) |
| Mental and behavorial disorders | 131 | 0.75 (0.62-0.89) | 1.05 (0.88-1.24) | 0.89 (0.75-1.07) | 17 | 0.81 (0.47-1.30) | 1.22 (0.71-1.96) | 0.97 (0.55-1.53) |
| Alcohol abuse | 92 | 0.85 (0.68-1.04) | 1.01 (0.81-1.24) | 1.02 (0.82-1.26) | 3 | 0.64 (0.13-1.88) | 0.70 (0.14-2.04) | 0.77 (0.16-2.21) |
| Diseases of the nervous system | 226 | 0.95 (0.83-1.08) | 1.66 (1.45-1.89) | 1.14 (1.00-1.31) | 54 | 0.96 (0.72-1.25) | 1.51 (1.14-1.97) | 1.15 (0.84-1.49) |
| Parkinson's disease | 42 | 0.82 (0.59-1.11) | 1.52 (1.09-2.05) | 0.99 (0.71-1.34) | 5 | 0.72 (0.23-1.68) | 1.06 (0.35-2.48) | 0.86 (0.27-1.97) |
| Alzheimer's disease | 73 | 0.89 (0.70-1.12) | 1.84 (1.44-2.31) | 1.07 (0.84-1.35) | 41 | 1.24 (0.89-1.68) | 2.07 (1.49-2.81) | 1.49 (1.05-2.01) |
| Diseases of the circulatory system | 1216 | 0.84 (0.79-0.88) | 1.36 (1.28-1.44) | 1.00 (0.94-1.08) | 299 | 0.89 (0.79-0.99) | 1.31 (1.17-1.47) | 1.07 (0.92-1.22) |
| Ischemic heart diseases | 150 | 0.74 (0.63-0.87) | 1.13 (0.95-1.32) | 0.89 (0.75-1.05) | 34 | 0.92 (0.64-1.28) | 1.29 (0.89-1.80) | 1.10 (0.74-1.52) |
| Other heart diseases | 311 | 0.83 (0.74-0.93) | 1.40 (1.25-1.56) | 1.00 (0.89-1.13) | 70 | 0.82 (0.64-1.04) | 1.27 (0.99-1.60) | 0.99 (0.74-1.23) |
| Cerebrovascular diseases | 338 | 0.60 (0.54-0.67) | 0.95 (0.86-1.06) | 0.70 (0.63-0.79) | 70 | 0.56 (0.44-0.71) | 0.80 (0.62-1.01) | 0.67 (0.48-0.80) |
| Diseases of the respiratory system | 213 | 0.75 (0.66-0.86) | 1.37 (1.19-1.56) | 0.90 (0.78-1.04) | 30 | 0.59 (0.40-0.85) | 0.96 (0.65-1.38) | 0.71 (0.46-0.99) |
| Pneumonia | 85 | 0.73 (0.58-0.90) | 1.40 (1.12-1.73) | 0.87 (0.70-1.09) | 16 | 0.79 (0.45-1.29) | 1.33 (0.76-2.17) | 0.95 (0.53-1.51) |
| Chronic lower respiratory diseases | 63 | 0.81 (0.63-1.04) | 1.27 (0.97-1.62) | 0.98 (0.75-1.26) | 4 | 0.38 (0.10-0.97) | 0.53 (0.15-1.37) | 0.46 (0.12-1.14) |
| Diseases of the digestive system | 216 | 0.84 (0.73-0.96) | 1.21 (1.06-1.39) | 1.01 (0.88-1.17) | 36 | 0.76 (0.53-1.05) | 1.04 (0.73-1.44) | 0.91 (0.61-1.23) |
| Cirrhosis, fibrosis and chronic hepatitis | 75 | 0.84 (0.66-1.06) | 1.03 (0.81-1.29) | 1.01 (0.80-1.28) | 12 | 1.20 (0.62-2.09) | 1.33 (0.69-2.33) | 1.44 (0.72-2.47) |
| Diseases of the skin and subcutaneous tissues | 28 | 1.21 (0.81-1.75) | 2.07 (1.38-3.00) | 1.46 (0.97-2.12) | 3 | 0.42 (0.09-1.23) | 0.68 (0.14-1.99) | 0.50 (0.10-1.44) |
| Diseases of the musculoskeletal system/connective tissue | 12 | 0.67 (0.35-1.18) | 1.12 (0.58-1.96) | 0.81 (0.42-1.42) | 6 | 1.03 (0.38-2.23) | 1.43 (0.52-3.11) | 1.23 (0.44-2.63) |
| Diseases of the genitourinary system | 89 | 0.81 (0.65-1.00) | 1.39 (1.11-1.71) | 0.98 (0.79-1.21) | 14 | 0.73 (0.40-1.23) | 1.04 (0.57-1.75) | 0.88 (0.46-1.44) |
| Diseases of the kidney and ureter | 54 | 0.78 (0.59-1.02) | 1.26 (0.95-1.65) | 0.93 (0.70-1.23) | 10 | 0.68 (0.33-1.25) | 0.95 (0.46-1.75) | 0.82 (0.38-1.47) |
| Symptoms, signs, ill-defined causes | 313 | 0.78 (0.70-0.87) | 1.48 (1.32-1.66) | 0.93 (0.83-1.05) | 88 | 0.86 (0.69-1.06) | 1.50 (1.20-1.85) | 1.03 (0.80-1.26) |
| External causes | 276 | 0.81 (0.71-0.91) | 1.06 (0.94-1.19) | 0.96 (0.85-1.09) | 27 | 0.74 (0.49-1.08) | 1.02 (0.67-1.48) | 0.89 (0.57-1.27) |

**Supplementary table 3**. Standardized mortality ratios (SMRs), causal mortality ratios (CMRs) and relative SMRs (rSMRs) by employment status

| | Farm owners | | | | Farm workers | | | |
|---|---|---|---|---|---|---|---|---|
| | **Obs** | **SMR (95% CI)** | **CMR (CI 95%)** | **rSMR (95%CI)** | **Obs** | **SMR (95% CI)** | **CMR (CI 95%)** | **rSMR (95%CI)** |
| All causes of death | 3402 | 0.79 (0.77-0.82) | 1.30 (1.26-1.35) | | 1763 | 0.92 (0.88-0.97) | 1.32 (1.26-1.38) | |
| Infectious and parasitic diseases | 76 | 0.60 (0.47-0.75) | 0.95 (0.75-1.18) | 0.75 (0.59-0.94) | 51 | 0.77 (0.57-1.01) | 1.05 (0.78-1.38) | 0.83 (0.62-1.10) |
| All cancers | 924 | 0.84 (0.79-0.90) | 1.32 (1.23-1.41) | 1.08 (1.00-1.17) | 468 | 0.94 (0.86-1.03) | 1.29 (1.17-1.41) | 1.03 (0.92-1.14) |
| Lip, buccal cavity, pharynx | 52 | 0.99 (0.74-1.29) | 1.26 (0.94-1.65) | 1.25 (0.93-1.64) | 18 | 0.81 (0.48-1.28) | 0.97 (0.58-1.54) | 0.88 (0.52-1.39) |
| Esophagus | 37 | 0.81 (0.57-1.11) | 1.05 (0.74-1.45) | 1.02 (0.72-1.41) | 14 | 0.80 (0.44-1.35) | 0.98 (0.54-1.64) | 0.87 (0.47-1.46) |
| Stomach | 117 | 1.04 (0.86-1.25) | 1.56 (1.29-1.86) | 1.33 (1.09-1.60) | 57 | 1.21 (0.92-1.57) | 1.65 (1.25-2.14) | 1.32 (1.00-1.72) |
| Colon, rectum and anus | 59 | 0.86 (0.65-1.10) | 1.39 (1.06-1.80) | 1.08 (0.82-1.40) | 35 | 0.85 (0.59-1.19) | 1.18 (0.82-1.64) | 0.92 (0.64-1.29) |
| Liver | 16 | 0.38 (0.22-0.62) | 0.54 (0.31-0.87) | 0.48 (0.27-0.77) | 17 | 0.82 (0.47-1.30) | 1.04 (0.60-1.66) | 0.88 (0.51-1.42) |
| Pancreas | 42 | 0.81 (0.59-1.10) | 1.25 (0.90-1.69) | 0.98 (0.74-1.39) | 19 | 0.68 (0.41-1.06) | 0.91 (0.55-1.43) | 0.81 (0.44-1.15) |
| Larynx | 12 | 0.61 (0.31-1.06) | 0.82 (0.42-1.44) | 0.73 (0.40-1.34) | 7 | 1.04 (0.42-2.13) | 1.30 (0.52-2.67) | 1.24 (0.45-2.32) |
| Lung | 63 | 0.68 (0.52-0.86) | 0.96 (0.74-1.23) | 0.81 (0.65-1.09) | 43 | 0.99 (0.72-1.34) | 1.28 (0.93-1.73) | 1.19 (0.78-1.46) |
| Skin | 4 | 0.87 (0.24-2.22) | 1.25 (0.34-3.20) | 1.04 (0.30-2.79) | 0 | - | - | 0.00 (-) |
| Breast | 11 | 0.90 (0.45-1.61) | 1.20 (0.60-2.14) | 1.08 (0.57-2.03) | 16 | 0.78 (0.44-1.26) | 0.92 (0.53-1.50) | 0.93 (0.48-1.37) |
| Uterus | 12 | 1.20 (0.62-2.09) | 1.56 (0.81-2.72) | 1.44 (0.78-2.64) | 15 | 0.92 (0.51-1.51) | 1.09 (0.61-1.81) | 1.10 (0.56-1.64) |
| Ovary | 1 | 0.34 (0.01-1.90) | 0.45 (0.01-2.53) | 0.41 (0.01-2.40) | 4 | 0.75 (0.21-1.93) | 0.89 (0.24-2.28) | 0.90 (0.22-2.09) |
| Prostate | 284 | 0.92 (0.82-1.03) | 1.72 (1.53-1.93) | 1.11 (1.04-1.33) | 88 | 0.99 (0.79-1.22) | 1.72 (1.38-2.11) | 1.19 (0.86-1.34) |
| Kidney | 2 | 0.26 (0.03-0.94) | 0.36 (0.04-1.31) | 0.31 (0.04-1.18) | 2 | 0.50 (0.06-1.81) | 0.63 (0.08-2.26) | 0.60 (0.07-1.97) |
| Bladder | 20 | 0.94 (0.57-1.45) | 1.55 (0.95-2.39) | 1.13 (0.72-1.83) | 6 | 0.62 (0.23-1.36) | 0.90 (0.33-1.95) | 0.75 (0.25-1.47) |
| Brain and central nervous system | 6 | 0.58 (0.21-1.26) | 0.75 (0.28-1.63) | 0.69 (0.27-1.59) | 6 | 0.88 (0.32-1.90) | 1.03 (0.38-2.25) | 1.05 (0.35-2.07) |
| Lymphatic and hematopoietic tissues | 69 | 0.80 (0.62-1.01) | 1.25 (0.98-1.59) | 0.96 (0.78-1.28) | 47 | 1.00 (0.74-1.33) | 1.36 (1.00-1.81) | 1.21 (0.80-1.46) |
| Diseases of the blood and of blood-forming organs | 11 | 0.65 (0.33-1.17) | 1.05 (0.52-1.87) | 0.78 (0.41-1.47) | 6 | 0.67 (0.24-1.45) | 0.88 (0.32-1.92) | 0.80 (0.26-1.57) |
| Endocrine, nutritional and metabolic disease | 182 | 0.78 (0.67-0.90) | 1.35 (1.16-1.57) | 0.93 (0.84-1.14) | 105 | 0.97 (0.79-1.17) | 1.46 (1.20-1.77) | 1.16 (0.85-1.28) |
| Diabetes mellitus | 127 | 0.78 (0.65-0.93) | 1.31 (1.09-1.56) | 0.93 (0.82-1.17) | 89 | 1.15 (0.92-1.41) | 1.69 (1.36-2.08) | 1.39 (1.01-1.56) |

| | Farm owners | | | | Farm workers | | | |
|---|---|---|---|---|---|---|---|---|
| | Obs | SMR (95% CI) | CMR (CI 95%) | rSMR (95%CI) | Obs | SMR (95% CI) | CMR (CI 95%) | rSMR (95%CI) |
| Mental and behavorial disorders | 84 | 0.63 (0.50-0.78) | 0.92 (0.74-1.14) | 0.75 (0.62-0.98) | 64 | 1.02 (0.79-1.31) | 1.34 (1.03-1.71) | 1.23 (0.85-1.43) |
| Alcohol abuse | 48 | 0.64 (0.47-0.85) | 0.77 (0.57-1.02) | 0.76 (0.59-1.06) | 47 | 1.24 (0.91-1.65) | 1.41 (1.04-1.88) | 1.49 (0.99-1.81) |
| Diseases of the nervous system | 182 | 0.92 (0.79-1.07) | 1.67 (1.43-1.93) | 1.11 (1.00-1.36) | 98 | 1.01 (0.82-1.23) | 1.56 (1.26-1.90) | 1.21 (0.88-1.34) |
| Parkinson's disease | 30 | 0.70 (0.47-1.00) | 1.30 (0.88-1.86) | 0.84 (0.60-1.27) | 17 | 1.11 (0.65-1.78) | 1.81 (1.05-2.90) | 1.33 (0.70-1.94) |
| Alzheimer's disease | 71 | 0.94 (0.73-1.18) | 1.93 (1.51-2.43) | 1.13 (0.92-1.50) | 43 | 1.09 (0.79-1.46) | 1.89 (1.37-2.55) | 1.31 (0.85-1.60) |
| Diseases of the circulatory system | 1041 | 0.82 (0.77-0.87) | 1.36 (1.27-1.44) | 0.98 (0.97-1.12) | 474 | 0.91 (0.83-1.00) | 1.34 (1.22-1.46) | 1.10 (0.88-1.09) |
| Ischemic heart diseases | 127 | 0.77 (0.64-0.91) | 1.18 (0.99-1.41) | 0.92 (0.80-1.15) | 57 | 0.78 (0.59-1.02) | 1.09 (0.82-1.41) | 0.94 (0.64-1.10) |
| Other heart diseases | 270 | 0.82 (0.72-0.92) | 1.40 (1.24-1.58) | 0.98 (0.91-1.17) | 111 | 0.86 (0.71-1.04) | 1.31 (1.08-1.57) | 1.04 (0.76-1.13) |
| Cerebrovascular diseases | 280 | 0.57 (0.51-0.64) | 0.92 (0.81-1.03) | 0.67 (0.61-0.78) | 128 | 0.65 (0.55-0.78) | 0.94 (0.79-1.12) | 0.78 (0.57-0.82) |
| Diseases of the respiratory system | 184 | 0.76 (0.65-0.88) | 1.40 (1.21-1.62) | 0.91 (0.82-1.11) | 59 | 0.65 (0.49-0.83) | 1.06 (0.80-1.36) | 0.77 (0.52-0.90) |
| Pneumonia | 70 | 0.70 (0.55-0.88) | 1.36 (1.06-1.72) | 0.84 (0.68-1.11) | 31 | 0.85 (0.57-1.20) | 1.46 (0.99-2.07) | 1.01 (0.62-1.30) |
| Chronic lower respiratory diseases | 52 | 0.79 (0.59-1.04) | 1.25 (0.93-1.64) | 0.95 (0.74-1.31) | 15 | 0.68 (0.38-1.11) | 0.96 (0.53-1.58) | 0.81 (0.41-1.21) |
| Diseases of the digestive system | 170 | 0.81 (0.69-0.94) | 1.20 (1.03-1.39) | 0.97 (0.87-1.19) | 82 | 0.87 (0.69-1.08) | 1.16 (0.92-1.44) | 1.04 (0.74-1.17) |
| Cirrhosis, fibrosis and chronic hepatitis | 54 | 0.81 (0.61-1.06) | 1.00 (0.75-1.31) | 0.97 (0.76-1.33) | 33 | 1.03 (0.71-1.44) | 1.19 (0.82-1.67) | 1.23 (0.77-1.57) |
| Diseases of the skin and subcutaneous tissues | 23 | 1.08 (0.68-1.62) | 1.89 (1.20-2.83) | 1.30 (0.86-2.05) | 8 | 0.89 (0.39-1.76) | 1.40 (0.61-2.77) | 1.07 (0.42-1.91) |
| Diseases of the musculoskeletal system/connective tissue | 12 | 0.79 (0.41-1.38) | 1.35 (0.70-2.36) | 0.95 (0.51-1.74) | 6 | 0.71 (0.26-1.54) | 1.00 (0.37-2.18) | 0.85 (0.28-1.67) |
| Diseases of the genitourinary system | 62 | 0.67 (0.52-0.86) | 1.16 (0.89-1.49) | 0.81 (0.65-1.09) | 41 | 1.12 (0.80-1.52) | 1.69 (1.21-2.29) | 1.35 (0.87-1.66) |
| Diseases of the kidney and ureter | 40 | 0.68 (0.49-0.93) | 1.13 (0.80-1.53) | 0.82 (0.61-1.17) | 24 | 0.95 (0.61-1.42) | 1.36 (0.87-2.02) | 1.14 (0.66-1.54) |
| Symptoms, signs, ill-defined causes | 245 | 0.71 (0.62-0.80) | 1.40 (1.23-1.59) | 0.84 (0.77-1.00) | 156 | 1.00 (0.85-1.17) | 1.65 (1.40-1.93) | 1.21 (0.92-1.29) |
| External causes | 171 | 0.70 (0.60-0.81) | 0.97 (0.83-1.12) | 0.83 (0.74-1.02) | 132 | 0.99 (0.83-1.17) | 1.20 (1.00-1.42) | 1.19 (0.90-1.29) |

**Supplementary table 4.** Standardized mortality ratios (SMRs), causal mortality ratios (CMRs) and relative SMRs (rSMRs) by region

| | Guadeloupe | | | | Martinique | | | |
|---|---|---|---|---|---|---|---|---|
| | **Obs** | **SMR (95% CI)** | **CMR (CI 95%)** | **rSMR (95%CI)** | **Obs** | **SMR (95% CI)** | **CMR (CI 95%)** | **rSMR (95%CI)** |
| All causes of death | 1958 | 0.83 (0.79-0.87) | 1.33 (1.27-1.39) | | 3207 | 0.83 (0.81-0.86) | 1.29 (1.25-1.34) | |
| Infectious and parasitic diseases | 43 | 0.64 (0.46-0.86) | 0.96 (0.69-1.29) | 0.76 (0.55-1.03) | 84 | 0.67 (0.53-0.83) | 1.00 (0.80-1.24) | 0.79 (0.63- 0.98) |
| All cancers | 527 | 0.89 (0.82-0.97) | 1.39 (1.27-1.51) | 1.10 (1.00-1.22) | 865 | 0.86 (0.80-0.92) | 1.27 (1.18-1.35) | 1.04 (0.96-1.13) |
| Lip, buccal cavity, pharynx | 35 | 1.15 (0.80-1.60) | 1.48 (1.03-2.05) | 1.39 (0.97-1.94) | 35 | 0.79 (0.55-1.09) | 0.97 (0.68-1.35) | 0.94 (0.65-1.31) |
| Esophagus | 30 | 1.09 (0.73-1.55) | 1.43 (0.96-2.04) | 1.31 (0.88-1.88) | 21 | 0.59 (0.37-0.90) | 0.74 (0.46-1.13) | 0.70 (0.44-1.08) |
| Stomach | 61 | 1.03 (0.79-1.32) | 1.53 (1.17-1.96) | 1.25 (0.95-1.61) | 113 | 1.13 (0.93-1.36) | 1.62 (1.33-1.95) | 1.37 (1.12-1.65) |
| Colon, rectum and anus | 28 | 0.81 (0.54-1.18) | 1.31 (0.87-1.89) | 0.98 (0.65-1.42) | 66 | 0.87 (0.68-1.11) | 1.30 (1.01-1.66) | 1.05 (0.81-1.34) |
| Liver | 17 | 0.67 (0.39-1.07) | 0.92 (0.54-1.48) | 0.80 (0.46-1.28) | 16 | 0.43 (0.24-0.69) | 0.57 (0.33-0.93) | 0.51 (0.29-0.83) |
| Pancreas | 21 | 0.74 (0.46-1.14) | 1.15 (0.71-1.75) | 0.89 (0.55- 1.37) | 40 | 0.78 (0.56-1.06) | 1.11 (0.79-1.51) | 0.93 (0.66- 1.27) |
| Larynx | 9 | 0.78 (0.36-1.48) | 1.07 (0.49-2.03) | 0.82 (0.63-1.04) | 10 | 0.67 (0.32-1.24) | 0.86 (0.41-1.59) | 0.81 (0.39-1.48) |
| Lung | 58 | 1.06 (0.80-1.37) | 1.50 (1.14-1.94) | 1.27 (0.97-1.66) | 48 | 0.59 (0.43-0.78) | 0.79 (0.59-1.05) | 0.70 (0.51-0.93) |
| Skin | 2 | 0.82 (0.10-2.95) | 1.11 (0.13-4.01) | 0.98 (0.12-3.55) | 2 | 0.44 (0.05-1.60) | 0.61 (0.07-2.19) | 0.53 (0.06-1.91) |
| Breast | 7 | 0.92 (0.37-1.90) | 1.23 (0.49-2.53) | 1.11 (0.45-2.29) | 20 | 0.79 (0.48-1.22) | 0.96 (0.59-1.49) | 0.95 (0.58-1.47) |
| Uterus | 7 | 1.04 (0.42-2.15) | 1.35 (0.54-2.77) | 1.25 (0.50-2.60) | 20 | 1.02 (0.62-1.57) | 1.23 (0.75-1.91) | 1.22 (0.74-1.89) |
| Ovary | 2 | 1.06 (0.13-3.84) | 1.43 (0.17-5.16) | 1.28 (0.15- 4.63) | 3 | 0.47 (0.10-1.38) | 0.57 (0.12-1.65) | 0.57 (0.12- 1.65) |
| Prostate | 130 | 0.85 (0.71-1.01) | 1.61 (1.34-1.91) | 1.02 (0.85-1.22) | 242 | 0.99 (0.87-1.13) | 1.79 (1.57-2.03) | 1.20 (1.05-1.37) |
| Kidney | 2 | 0.38 (0.05-1.36) | 0.53 (0.06-1.90) | 0.45 (0.05-1.64) | 2 | 0.31 (0.04-1.13) | 0.41 (0.05-1.47) | 0.38 (0.05-1.36) |
| Bladder | 9 | 0.90 (0.41-1.71) | 1.45 (0.66-2.76) | 1.08 (0.50-2.06) | 17 | 0.81 (0.47-1.30) | 1.27 (0.74-2.03) | 0.97 (0.56-1.56) |
| Brain and central nervous system | 3 | 0.46 (0.09-1.34) | 0.60 (0.12-1.75) | 0.55 (0.11-1.61) | 9 | 0.84 (0.39-1.60) | 1.02 (0.47-1.94) | 1.01 (0.46-1.92) |
| Lymphatic and hematopoietic tissues | 38 | 0.91 (0.64-1.25) | 1.42 (1.01-1.95) | 1.09 (0.77-1.51) | 78 | 0.85 (0.67-1.07) | 1.24 (0.98-1.55) | 1.02 (0.81-1.28) |
| Diseases of the blood and of blood-forming organs | 9 | 0.97 (0.45-1.85) | 1.58 (0.72-3.00) | 1.17 (0.54-2.23) | 8 | 0.48 (0.21-0.95) | 0.70 (0.30-1.37) | 0.58 (0.25-1.13) |
| Endocrine, nutritional and metabolic disease | 87 | 0.70 (0.56-0.87) | 1.20 (0.96-1.48) | 0.84 (0.67- 1.04) | 200 | 0.91 (0.79-1.05) | 1.49 (1.29-1.72) | 1.10 (0.95- 1.27) |
| Diabetes mellitus | 67 | 0.75 (0.58-0.95) | 1.24 (0.96-1.57) | 0.90 (0.69-1.15) | 149 | 0.99 (0.83-1.16) | 1.56 (1.32-1.83) | 1.19 (1.00-1.40) |

| | Guadeloupe | | | | Martinique | | | |
|---|---|---|---|---|---|---|---|---|
| | **Obs** | **SMR (95% CI)** | **CMR (CI 95%)** | **rSMR (95%CI)** | **Obs** | **SMR (95% CI)** | **CMR (CI 95%)** | **rSMR (95%CI)** |
| Mental and behavorial disorders | 57 | 0.75 (0.57-0.98) | 1.08 (0.82-1.40) | 0.90 (0.68-1.18) | 91 | 0.75 (0.61-0.92) | 1.06 (0.85-1.30) | 0.90 (0.72-1.11) |
| Alcohol abuse | 36 | 0.79 (0.55-1.09) | 0.96 (0.67-1.33) | 0.95 (0.66-1.32) | 59 | 0.87 (0.66-1.13) | 1.02 (0.77-1.31) | 1.05 (0.79-1.35) |
| Diseases of the nervous system | 110 | 1.11 (0.91-1.34) | 1.94 (1.59-2.33) | 1.44 (1.19-1.77) | 170 | 0.87 (0.74-1.01) | 1.47 (1.26-1.71) | 1.04 (0.89-1.22) |
| Parkinson's disease | 24 | 1.03 (0.66-1.54) | 2.00 (1.28-2.98) | 1.24 (0.80-1.86) | 23 | 0.66 (0.42-0.99) | 1.13 (0.71-1.69) | 0.79 (0.50-1.19) |
| Alzheimer's disease | 35 | 1.13 (0.79-1.57) | 2.32 (1.61-3.22) | 1.36 (0.95-1.91) | 79 | 0.94 (0.74-1.17) | 1.78 (1.41-2.22) | 1.13 (0.89-1.41) |
| Diseases of the circulatory system | 594 | 0.84 (0.77-0.91) | 1.37 (1.26-1.48) | 1.01 (0.92- 1.11) | 921 | 0.85 (0.80-0.91) | 1.34 (1.25-1.43) | 1.03 (0.95- 1.11) |
| Ischemic heart diseases | 78 | 0.83 (0.66-1.04) | 1.26 (1.00-1.57) | 1.00 (0.79-1.26) | 106 | 0.73 (0.60-0.88) | 1.08 (0.89-1.31) | 0.87 (0.71-1.06) |
| Other heart diseases | 163 | 0.85 (0.73-0.99) | 1.43 (1.22-1.67) | 1.02 (0.87-1.21) | 218 | 0.82 (0.71-0.93) | 1.33 (1.16-1.52) | 0.98 (0.85-1.12) |
| Cerebrovascular diseases | 166 | 0.61 (0.52-0.71) | 0.97 (0.83-1.13) | 0.72 (0.60-0.83) | 242 | 0.58 (0.51-0.66) | 0.90 (0.79-1.02) | 0.69 (0.59-0.77) |
| Diseases of the respiratory system | 101 | 0.82 (0.67-0.99) | 1.46 (1.19-1.78) | 0.98 (0.80-1.20) | 142 | 0.68 (0.57-0.80) | 1.20 (1.01-1.42) | 0.81 (0.67-0.95) |
| Pneumonia | 27 | 0.56 (0.37-0.81) | 1.04 (0.69-1.52) | 0.67 (0.44-0.97) | 74 | 0.84 (0.66-1.05) | 1.58 (1.24-1.98) | 1.01 (0.79-1.26) |
| Chronic lower respiratory diseases | 38 | 0.99 (0.70-1.35) | 1.56 (1.10-2.14) | 1.18 (0.84-1.64) | 29 | 0.59 (0.39-0.84) | 0.88 (0.59-1.26) | 0.70 (0.47-1.01) |
| Diseases of the digestive system | 94 | 0.77 (0.62-0.94) | 1.11 (0.89-1.35) | 0.92 (0.74- 1.14) | 158 | 0.87 (0.74-1.01) | 1.24 (1.05-1.45) | 1.04 (0.88- 1.22) |
| Cirrhosis, fibrosis and chronic hepatitis | 38 | 0.87 (0.61-1.19) | 1.07 (0.76-1.47) | 1.04 (0.74-1.44) | 49 | 0.89 (0.66-1.18) | 1.06 (0.79-1.41) | 1.07 (0.79-1.41) |
| Diseases of the skin and subcutaneous tissues | 11 | 1.14 (0.57-2.03) | 2.00 (1.00-3.58) | 1.36 (0.68-2.46) | 20 | 0.97 (0.59-1.50) | 1.63 (0.99-2.51) | 1.17 (0.71-1.80) |
| Diseases of the musculoskeletal system/connective tissue | 7 | 0.92 (0.37-1.90) | 1.52 (0.61-3.14) | 1.10 (0.44-2.29) | 11 | 0.68 (0.34-1.23) | 1.08 (0.54-1.93) | 0.82 (0.41-1.47) |
| Diseases of the genitourinary system | 41 | 0.83 (0.60-1.13) | 1.39 (1.00-1.89) | 1.00 (0.71-1.36) | 62 | 0.78 (0.60-1.00) | 1.29 (0.99-1.65) | 0.94 (0.72-1.21) |
| Diseases of the kidney and ureter | 33 | 0.98 (0.67-1.37) | 1.59 (1.09-2.23) | 1.17 (0.81-1.66) | 31 | 0.62 (0.42-0.88) | 0.96 (0.65-1.36) | 0.74 (0.50-1.05) |
| Symptoms, signs, ill-defined causes | 150 | 0.79 (0.67-0.92) | 1.55 (1.31-1.81) | 0.94 (0.79-1.12) | 251 | 0.80 (0.71-0.91) | 1.45 (1.28-1.65) | 0.96 (0.84-1.09) |
| External causes | 114 | 0.70 (0.58-0.85) | 0.94 (0.78-1.13) | 0.84 (0.69-1.01) | 189 | 0.87 (0.75-1.00) | 1.14 (0.98-1.31) | 1.05 (0.90-1.21) |
